# Symptoms associated with a COVID-19 infection in the general population of Vienna

**DOI:** 10.1101/2021.02.24.21252426

**Authors:** Nicolas Munsch, Stefanie Gruarin, Jama Nateqi, Thomas Lutz, Michael Binder, Judith H. Aberle, Alistair Martin, Bernhard Knapp

## Abstract

**Background:** Most clinical studies report the symptoms experienced by those infected with Coronavirus disease 2019 (COVID-19) via patients already hospitalised. Here we analyse the symptoms experienced by the general population in Vienna.

**Methods:** The Vienna Social Fund (FSW, Vienna, Austria), the Public Health Services of the City of Vienna (MA15) and the private company Symptoma collaborated to implement Vienna’s official online COVID-19 symptom checker. Users answered 12 yes/no questions about symptoms to assess their risk for COVID-19. They could also specify their age and sex, and whether they had contact with someone who tested positive for COVID-19. Depending on the assessed risk of COVID-19 positivity, a SARS-CoV-2 nucleic acid amplification test (NAAT) was performed. In this publication, we analysed which factors (symptoms, sex or age) are associated with COVID-19 positivity. We also trained a classifier to correctly predict COVID-19 positivity from the collected data.

**Results:** Between the 2nd of November 2020 and the 18th of November 2021, 9133 people experiencing COVID-19-like symptoms were assessed as high risk by the chatbot and were subsequently tested by a NAAT. Symptoms significantly associated with a positive COVID-19 test were malaise, fatigue, headache, cough, fever, dysgeusia and hyposmia. Our classifier could successfully predict COVID-19 positivity with an Area Under the Curve (AUC) of 0.74.

**Conclusion:** This study provides reliable COVID-19 symptom statistics based on the general population verified by NAATs.

## Introduction

The frequency of the symptoms associated with COVID-19 is valuable information for health authorities during the pandemic. Such knowledge has been used in a variety of applications including triage recommendation and diagnostics [1–3]. However, most studies reporting on COVID-19 symptom frequencies concern patients in hospital settings [1]. Thus, published symptom frequencies may not reflect those found in the general population. For example, non-hospitalised people experience less severe forms of the disease [1].

To reduce the above sampling bias, some approaches have been implemented to collect COVID-19 self-reported symptoms from the general population. Self-reported symptoms have been collected via a variety of means, including an official test prioritisation questionnaire [4], symptom trackers [5–7], a symptom checker [8], and, even, Twitter [9]. However, only the symptoms collected via an official test prioritization questionnaire are associated with verified tests results. With other methods, test results are only self-reported. Of the data associated with a verified result, while valuable, most have shortcomings. For example, that released by the Israeli Ministry of Health does not contain dysgeusia nor anosmia as it was not known as a relevant symptom at the early stages of the pandemic [4].

In this work, we describe the COVID-19 symptoms reported by the non-hospitalized Viennese population from the 2nd of November 2020 to the 18th of November 2021. With a dataset spanning over 9000 users, we analyse the association of these symptoms with a positive COVID-19 NAAT and build a classifier to predict COVID-19 positivity from those experiencing flu-like symptoms.

## Methods

### Data collection

From November 2020, Vienna’s online COVID-19 symptom checker provided inhabitants with an initial COVID-19 risk assessment. Depending on the outcome, possible options for further action included a NAAT (Nucleic Acid Amplification Test) using the Reverse transcription-polymerase chain reaction (RT-PCR) method [10,11]. The aim was to offer an additional scalable service, complementing the medical telephone health service “1450”. The symptom checker is currently available at https://symptomchecker.fsw.at/.

The Vienna Social Fund (FSW), the Public Health Services of the City of Vienna (MA15), and the private company Symptoma mutually developed the chatbot based on previous results detailing the accuracy of Symptoma’s symptom checker with regards to COVID-19 [3,12,13]. During the chatbot conversation, each user was asked the same set of questions and responses were recorded accordingly. A user had to answer a series of 12 yes/no questions about symptoms. These are fever (>38°C), cough, dyspnea, sneezing, rhinorrhea, sore throat, malaise, fatigue, diarrhea, headache, hyposmia, dysgeusia. In addition, the user could indicate if, in the last 10 days, there was close contact with a duration longer than 15 minutes with a person who tested positive for COVID-19. Finally, each user was invited to specify their age and sex. We did not record the exact age of users for data protection issues, but only the age group (see Supplementary Table 1). If the chatbot assessed a user to have a high risk for a COVID-19 infection [3], the user reported a positive self-test, the user returned from abroad, or the user had a severe medical precondition and any type of symptom then the user was offered a NAAT [10,11].

The statistics reported in this paper are based on the combined information of the chatbot conversations and the results of the NAATs. A total of 10089 users were screened this way between the 2nd of November 2020 and the 18th of November 2021. Among them, 956 did not report any symptoms but only close contact with a person who tested positive for COVID-19. These were excluded from our further analyses. The remaining 9133 users were used in the further analyses below.

### Data analysis

All data were anonymised prior to analysis. Only sex, age group, the answers to the questions, and the result of the NAAT were collated. We analysed for each symptom if there was a significant difference between users who tested positive for COVID-19 (C19+) and users who tested negative for COVID-19 (C19-). We repeated the analysis between males and females that are C19+, and between males and females that are C19-. P-values were calculated by a two-tailed Fisher’s exact test and corrected for multiple testing by the Benjamini-Hochberg [14] method. In addition, we quantified the association between symptoms via a two-tailed Fisher’s exact test. Log odds ratios (LOR) were calculated and P-values were corrected for multiple testing by the Benjamini-Hochberg [14] method. Lastly, we built a logistic regression model to predict C19+ based on the collected data. 8966 users, who provided an age group and specified their sex as either male or female, were included in this analysis. Performance was assessed based on the concatenation of the 10 test sets obtained from the cross-validation. We analysed the Receiving Operating Characteristics (ROC) curve and the area under the ROC curve (AUC). Confidence intervals (CI) were calculated by bootstrapping with 3000 repetitions. The same methodology was applied to train and evaluate other classifiers including Random Forest, AdaBoost and Support Vector Machine.

All analyses were done in Python 3.8 using the libraries Numpy (1.21.0) [15], Pandas (1.3.4) [16], Scikit-learn (1.0.1), and Statsmodels (0.13.1) [17]. Visualisations were produced using Matplotlib (3.5.1) [18] and Seaborn (0.11.2) [19].

## Results

### Symptom frequencies among COVID-19 positive and COVID-19 negative users

Summary statistics of participants and numerical details are given in Supplementary Table 1. Our study cohort consists of 9133 non-hospitalised persons experiencing flu-like symptoms. Of which, 2692 (29.5%) tested positive for COVID-19 (C19+) and 6341 (69.4%) tested negative for COVID-19 (C19-). The test was unclear for 100 persons (1.1%). The median group age is 31-40 years for C19+ and 21-30 for the C19-groups. In Figure 1, we compared the symptom frequencies between C19+ and C19-. The symptoms most frequently reported by C19+ users were malaise (78.6%), fatigue (73.8%), headache (63.7%), cough (59.8%), and fever (49.7%). Users less frequently reported sore throat (47.8%), close contact with a person who tested positive for COVID-19 (40.7%), rhinorrhea (38.1%), sneezing (33.9%), dysgeusia (28.9%), and hyposmia (26.0%). Dyspnea (15.0%) and diarrhea (11.8%) were rarely reported.

**Figure 1.**
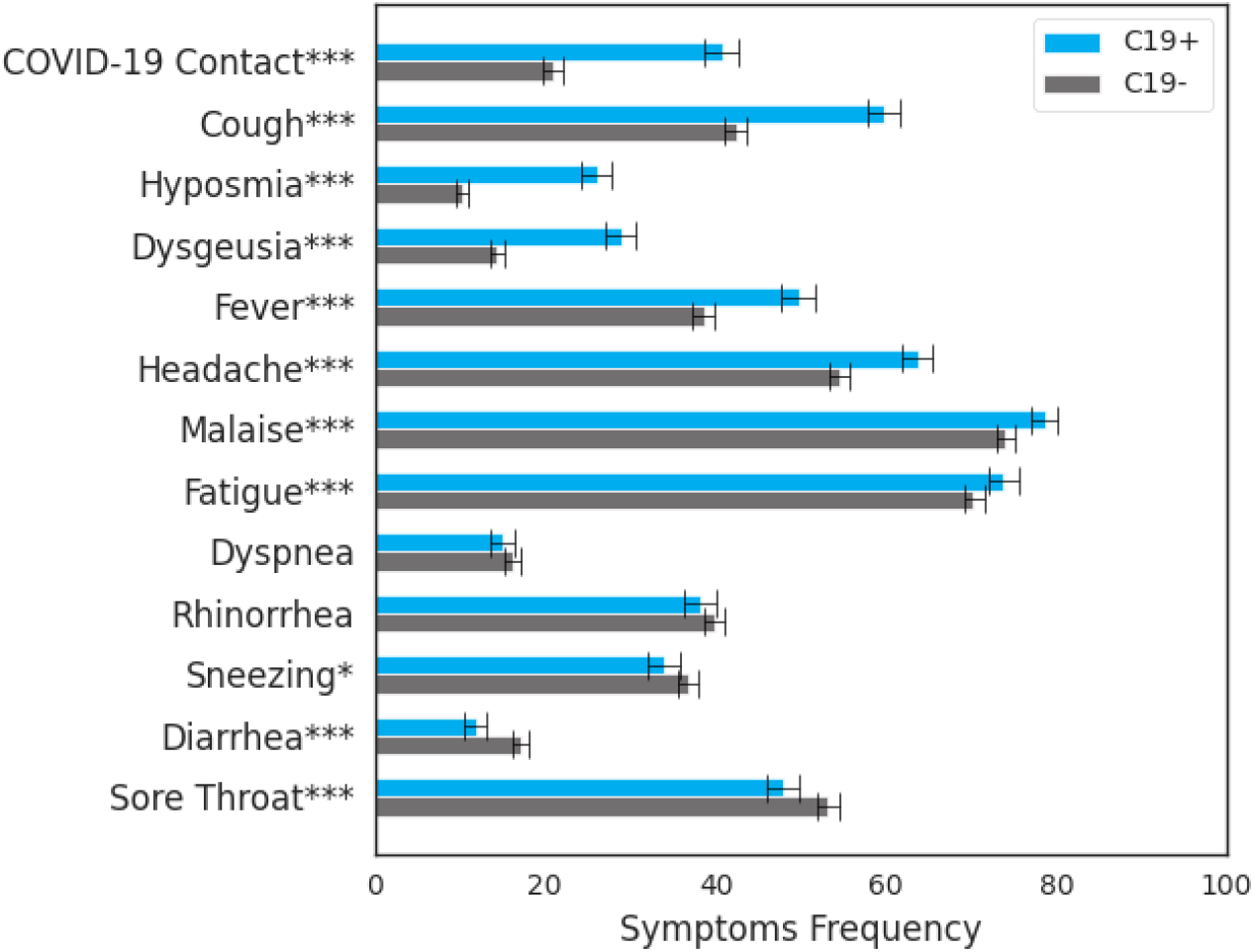
Symptom frequencies for the C19+ and C19-groups. Error bars indicate the 95% confidence intervals. Significance of the difference between these groups are indicated with one, two, and three asterisks which correspond to a p-value less than 0.05, 0.01, and 0.001 respectively.

C19+ users significantly more frequently reported cough (*P* < 0.001), hyposmia (*P* < 0.001), fever (*P* < 0.001), dysgeusia (*P* < 0.001), headache (*P* < 0.001), malaise (*P* < 0.001), fatigue (*P* < 0.001), and close contact with a person who tested positive for COVID-19 (*P* < 0.001). On the contrary, C19+ users significantly less frequently reported diarrhea (*P* < 0.001), sore throat (*P* < 0.001) and sneezing (*P* = 0.01). However, no significant difference between the C19+ and C19-groups was found for rhinorrhea (*P* = 0.12) and dyspnea (*P* = 0.17).

The largest increase in C19+ persons was found for close contact with a person who tested positive for COVID-19 (+19.8%), hyposmia (+15.8%) and dysgeusia (+14.1%). The largest decrease in C19+ persons was found for sore throat (−5.4%) and diarrhea (−5.3%).

In both C19+ and C19-groups, women reported sore throat more frequently (*P* < 0.001), sneezing (*P* < 0.001) and headache (*P* < 0.01) than men. Men reported fever more frequently (*P* < 0.001). In the C19+ group only, rhinorrhea and dyspnea (*P* < 0.01) are more frequently present for women than for men. In the C19-group only, men reported diarrhea more frequently (*P* = 0.02), while women more frequently reported fatigue (*P* < 0.001) (Supplementary Figure 1).

### Co-occurrence and association of symptoms

The frequency and the association of all pairs of symptoms within C19+ are indicated in Figure 2. For most of the pairs of symptoms (66.7%), the association is significantly positive. The three highest associations of symptoms within the C19+ group were between dysgeusia and hyposmia (LOR (Log Odds Ratio)=3.5, *P* < 0.001), fatigue and malaise (LOR=2.2, *P* < 0.001), and sneezing and rhinorrhea (LOR=1.9, *P* < 0.001). Within the C19+ group, 21% reported both dysgeusia and hyposmia, 23% reported both rhinorrhea and sneezing and 66% reported both fatigue and malaise. These associations were also observed within the C19-group (see Supplementary Figure 2.) Among associations with a LOR higher than 0.15, only those between fever and diarrhea (LOR=0.25, *P* = 0.052), malaise and dysgeusia (LOR=0.20, *P* = 0.083), and dysgeusia and headache (LOR=0.18, *P* = 0.063) are not significant. Associations with a LOR between -0.15 and 0.15 don’t have a significant positive or negative association.

**Figure 2.**
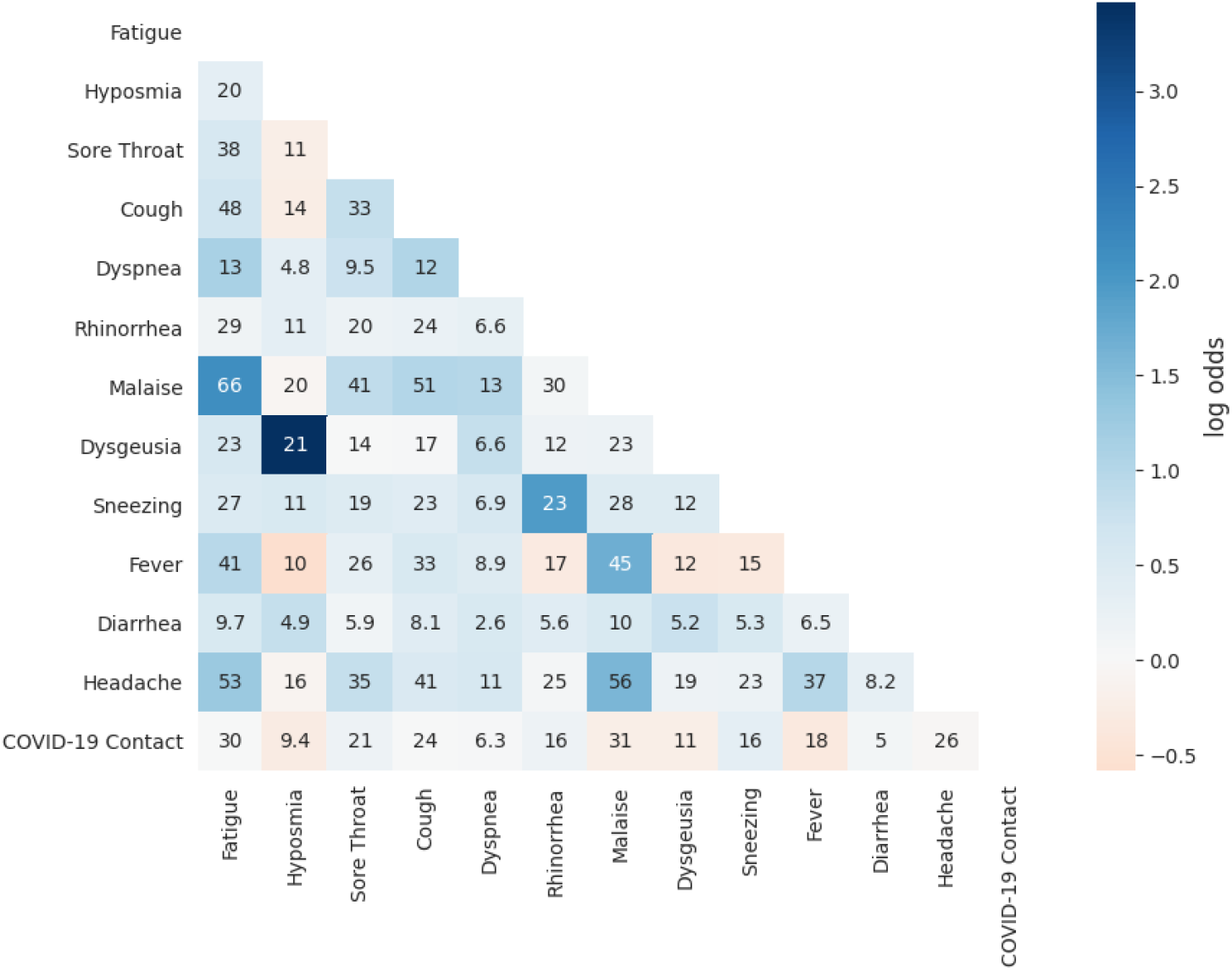
Symptoms co-occurrence frequencies for the C19+ group. Frequencies are reported in percentage. Log Odds Ratios (LOR) are represented by the colour scale. They show the strength of the association. LOR indicates an association when its value is more than 1, a dissociation if lower than 1. The equivalent results for the C19-group is included as Supplementary Figure 2.

The three strongest significantly negative associations were between fever and hyposmia (LOR=-0.58, *P* < 0.001), fever and dysgeusia (LOR=-0.37, *P* < 0.001), and fever and sneezing (LOR=-0.34, *P* < 0.001).

### Evaluation of a classifier based on symptoms

A classifier was built using the 8966 users that reported symptoms, age, and sex. The ROCs for each fold are shown in Figure 3. Across the 10 folds, the logistic regression model predicts with an AUC of 0.74 [0.72, 0.75]. Similar performances were obtained by the Random Forest, AdaBoost and SVM models. Possible working points, that being a threshold to which we predict COVID-19 positivity, include a sensitivity of 0.70 and a specificity of 0.65, sensitivity of 0.80 and a specificity of 0.51, or a sensitivity of 0.90 and a specificity of 0.32. We also evaluated the performance when excluding the answer about a Contact with a COVID-19 positively tested person. In this setting, the AUC is 0.69 [0.68. 0.70]. After training on all the available data, the following equation was found for the logistic regression:

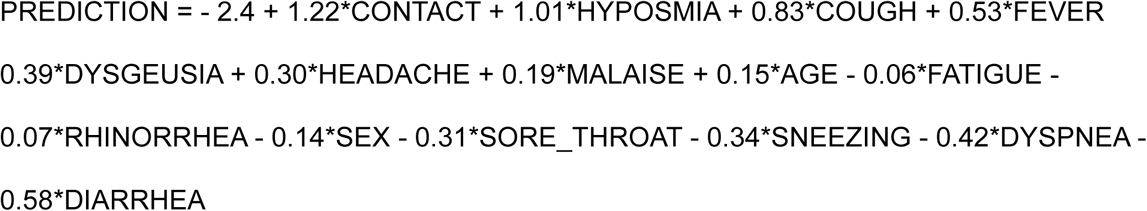

**Fig 3.**
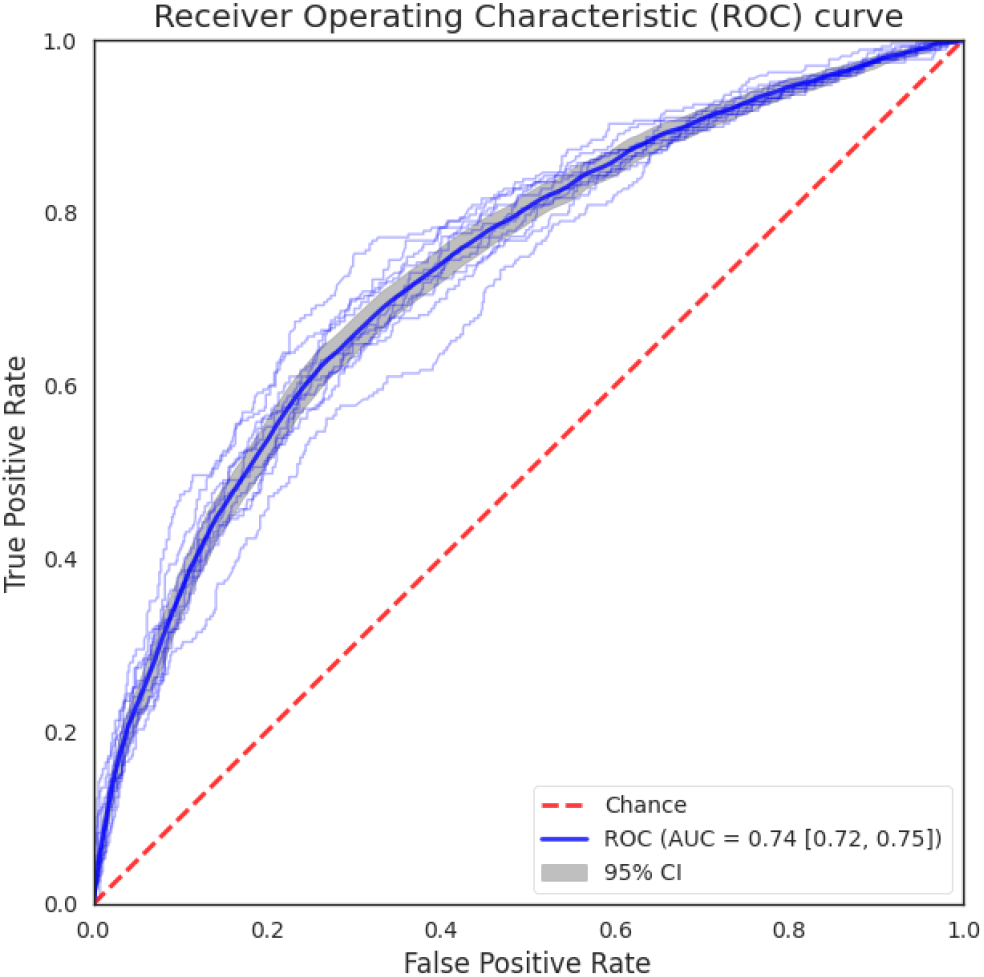
Receiver operating characteristic curve (ROC) of the Logistic Regression model when accounting for the contact with COVID-19 case information. The transparent band shows the 95% Confidence Intervals (CI). The area under the curve (AUC) is provided to summarize the curve. The ROCs for the Random Forest, AdaBoost and Support Vector Machine models are provided in Supplementary Figure 3. An alternative version of the ROC for the Logistic Regression model without using the contact with COVID-19 case information is included in Supplementary Figure 4.

Sex information is encoded to be 1 for the female and 0 for the male. Age categories were encoded as integers.

## Discussion

To the best of our knowledge, this is the first study reporting symptoms associated with COVID-19 of the general population presenting symptoms, tested by NAATs. Our results showed a cough frequency of 59.8% which falls within the 95% CI of the symptom frequencies reported in literature (range [59.8%-74.1%]) [1]. Similar agreements were found for hyposmia (26.0% vs [17.7%-41.3%]), dysgeusia (28.9% vs [12.4%-43.5%]), diarrhea (11.8% vs [7.6%-17.4%]) and fever (49.7% vs [35.0%-71.7%]). The frequency of dyspnea (15.0%) was slightly below the 95% CI reported (16.6%-35.5%), while the frequencies of headache (63.7% vs [9.2%-43.5%]), fatigue (73.8% vs [22.1%-53.6%]) and sore throat (47.8% vs [13.5%-31.6%]) were higher [1]. This discrepancy might be explained by the difference in interpreting the symptoms between patient and physician [21]. For example, fatigue was questioned by the associated chatbot, as “Have you been feeling particularly fatigued or dull lately?” (in german: “Fühlen Sie sich neuerdings besonders ermüdet oder matt?”).

The results also show that the symptoms experienced by C19+ significantly differ, except for dyspnea, to those experienced by C19-. This agrees with other studies that also reported hyposmia, dysgeusia, and fever as significantly increased in C19+ persons [6,22,23]. Further, the large relative difference of hyposmia and dysgeusia frequencies for C19+ users suggests that hyposmia and dysgeusia are specific but not sensitive, i.e. their presence strongly suggests the user is C19+ but no conclusion can be drawn from their absence.

The association of dyspnea with a COVID-19 positivity, not observed in the present analysis, was also not found by Menni et al. [2]. This might be due to a sample bias as dyspnea is often a late symptom of an infection while chatbot users might rather be at an earlier stage of infection [24]. Alternatively, dyspnea can be a distressing symptom and affected individuals might rather call an emergency hotline instead of using a chatbot [25]. Additionally, results show symptoms were reported in different frequencies by men and women, which could be caused by sex-specific differences in the clinical course [26,27].

As shown in previous studies, the high correlation found between dysgeusia and hyposmia indicates that these pairs of symptoms frequently occur together [28]. The same holds true for rhinorrhea and sneezing [29]. The high correlation between fatigue and malaise might be explained by the fact that fatigue is a subjective symptom of malaise [30]. The latter pair of symptoms also has a high co-occurrence frequency, which might be explained by the correlation, and the non-specific nature of these symptoms [30].

The AUC of our predictor (0.74) is in the range of the performance of the symptom-based COVID-19 predictor described in the literature. Other reported AUCs were as 0.68 [22], 0.74 [2] and 0.90 [4]. The considerably higher AUC of the latter predictor is explained by the inclusion of many asymptomatic patients who did not report any contact with a COVID-19 infected person. These patients, as expected, are mostly C19-, thereby inflating performance. Predicting COVID-19 positivity from patients who don’t report any symptoms or contact is not considered within our study, which only deals with symptomatic people.

Our study has limitations. First, self-reported symptoms are, by definition, not assessed by a medical professional which leads to inconsistencies. Second, there is selection bias because people with a low risk of being C19+ were not offered a test (see methods). Another selection bias is that chatbot usage is correlated with age (see Supplementary Table 1 and Fagherazzi et al. [31]). Third, users can experience additional symptoms after completing the session. These symptoms were not recorded and included in the present study. This leads us to believe, as discussed previously, that our cohort is focused on the early onset of COVID-19. Lastly, the consideration of the NAAT as a ground truth has been criticised due to its low sensitivity [32].

In addition to the above limitations, our sampling period aligns with Austria’s COVID-19 vaccination campaign as well as the emergence of new variants. Both of these factors, namely being vaccinated or being infected with a variant, have the potential to alter the symptoms experienced. For example, it has been reported that vaccinations reduce the number of symptoms experienced [33]. In contrast, the emergence of variant B.1.1.7 (Alpha), was reported to not affect the symptoms experienced [34]. In our study, we did not find any significant changes in the symptom frequencies over time (see Supplementary Figure 5).

In conclusion, we have analysed in-depth the COVID-19 symptoms reported by the non-hospitalized Viennese population over the past year. Data were systematically collected and results were automatically associated with a NAAT. To date, no other work features a general European population in combination with such rigorous data collection. For this reason, we believe that this work provides excellent new insights into the characteristics of COVID-19, both alone and when compared to other flu-like diseases.

## Data Availability

All relevant data is contained within the manuscript

## Data Availability

All relevant data is reported within the study.

## Acknowledgement

We would like to thank the Vienna Social Fund (FSW), Public Health Services of the City of Vienna (MA15) and the Vienna Health Authority for providing data and making this publication possible.

## Ethical considerations

This study was exempted from ethics review by the ethics commission of the city of Vienna (MA15-EK/21-037-VK_NZ). All individuals using the chatbot agreed that their data will be used in an anonymised way.

## Author contributions

Study design: BK, JN, TL. Data compilation: NM, TL, MB. Data analysis and critical revision: NM, BK, AM, JA. Writing the manuscript: NM, BK. Revising the manuscript critically: BK, SG, AM, JN, JA, MB.

---

**Supplementary Table 1:**
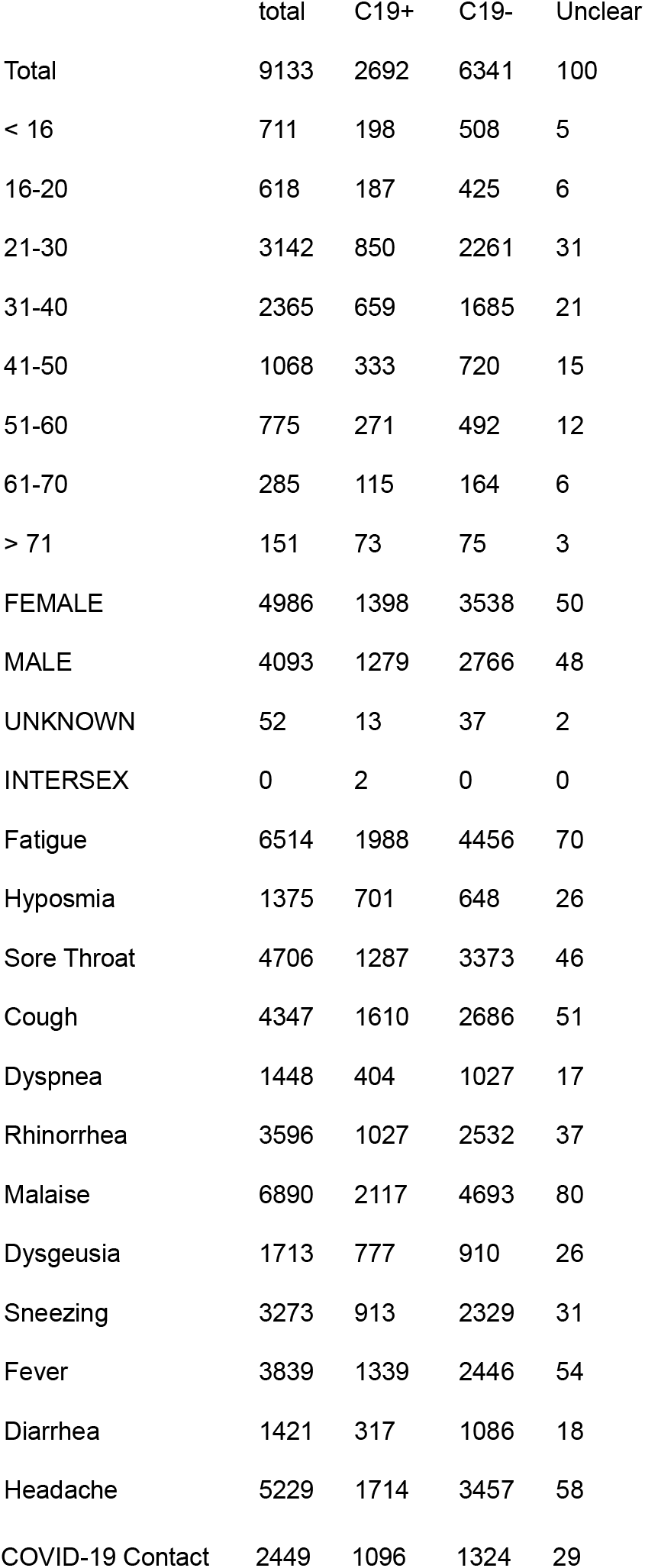
Users characteristics

**Supplementary Figure 1.**
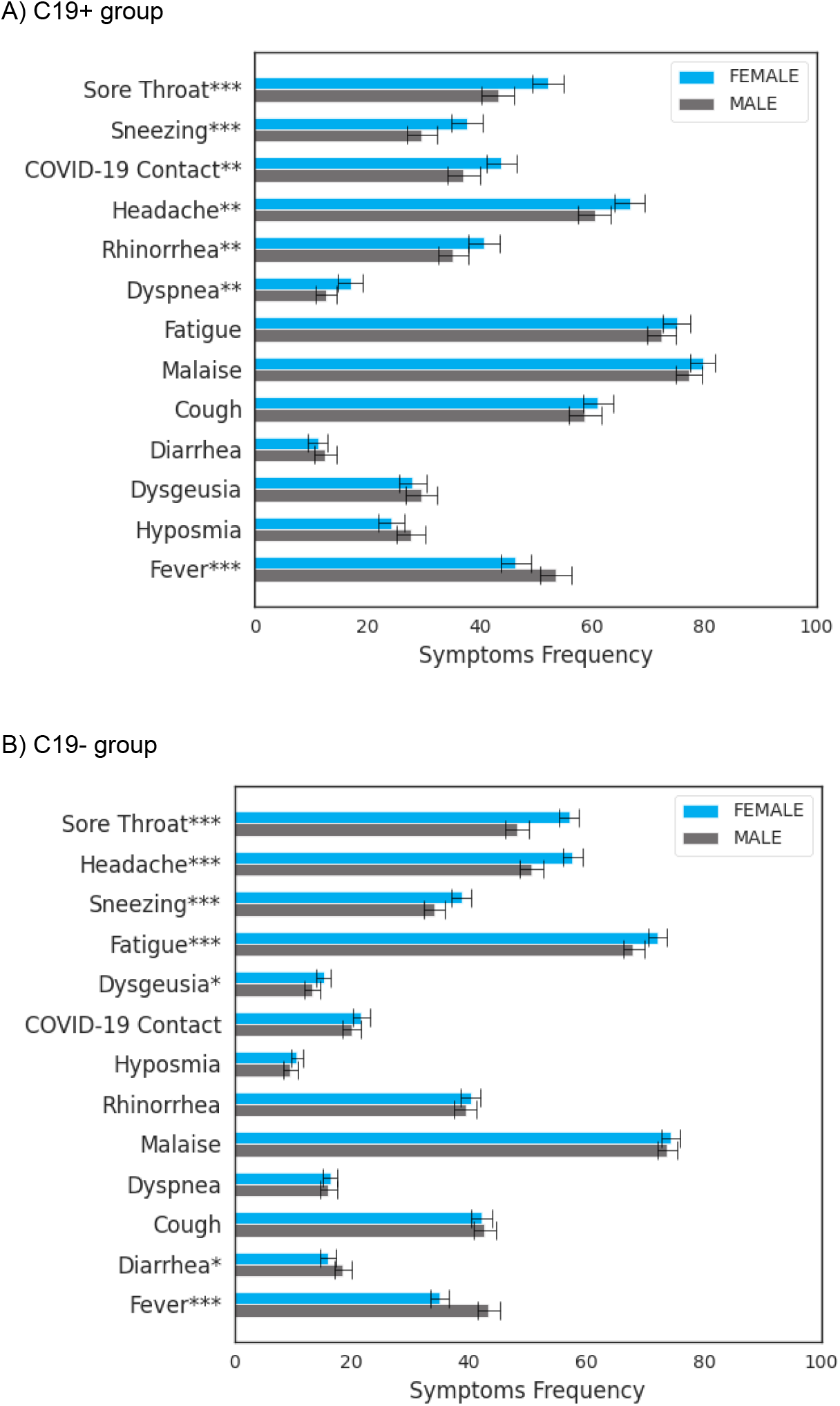
Symptom frequencies among the female and male for the C19+ group and (B) for the C19-group. Error bars show the 95% confidence intervals. Symptoms with a p-value less than 0.05, 0.01, and 0.001 are indicated with one, two, and three asterisks respectively.

**Supplementary Figure 2.**
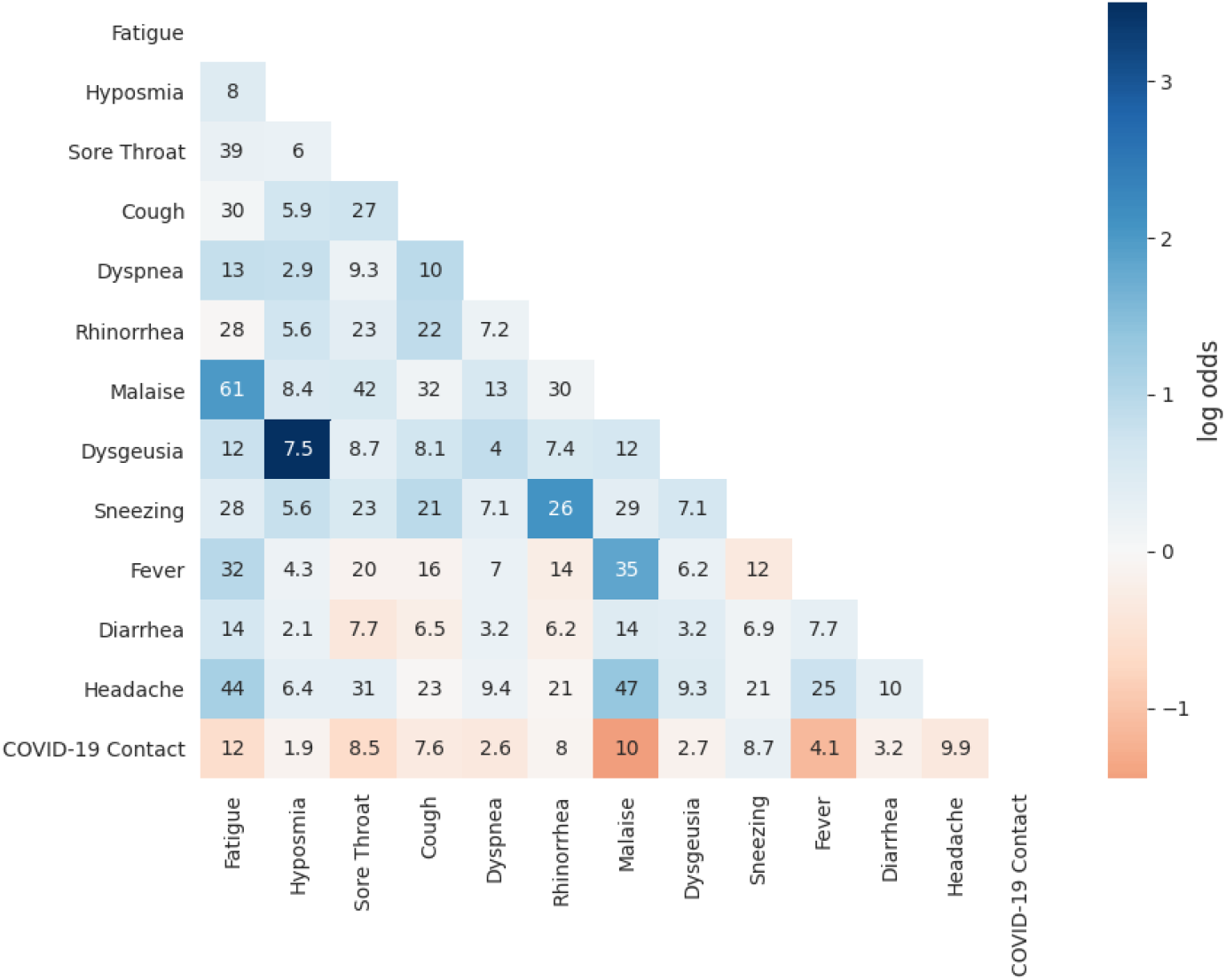
Symptoms co-occurrence frequencies for the C19-group. Frequencies are reported in percentage. Log Odds Ratios (LOR) are represented by the color scale. They show the strength of the association. LOR indicates a strong association when its value is more than 1, an inverted association if lower than 1 and no association if close to 1.

**Supplementary Figure 4.**
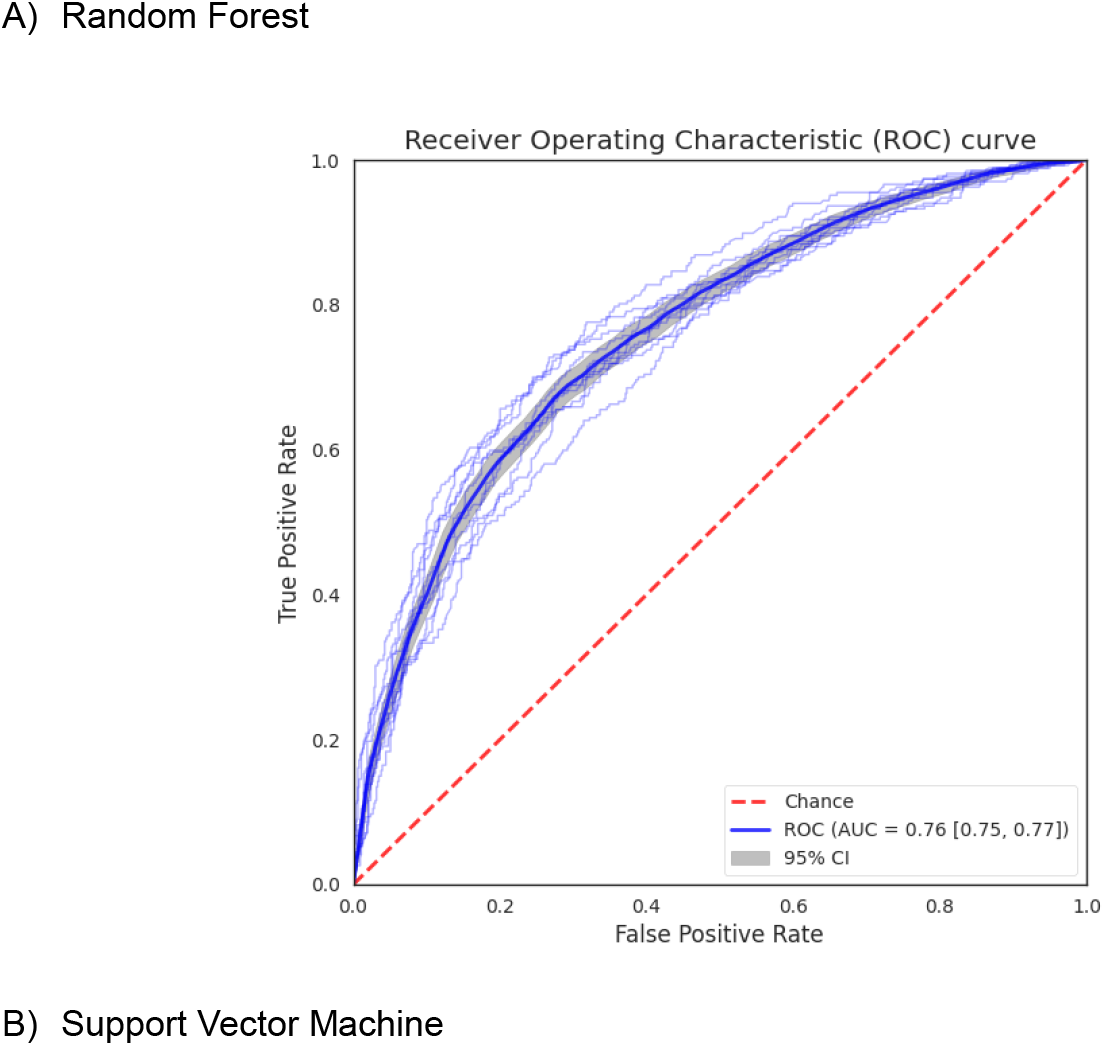
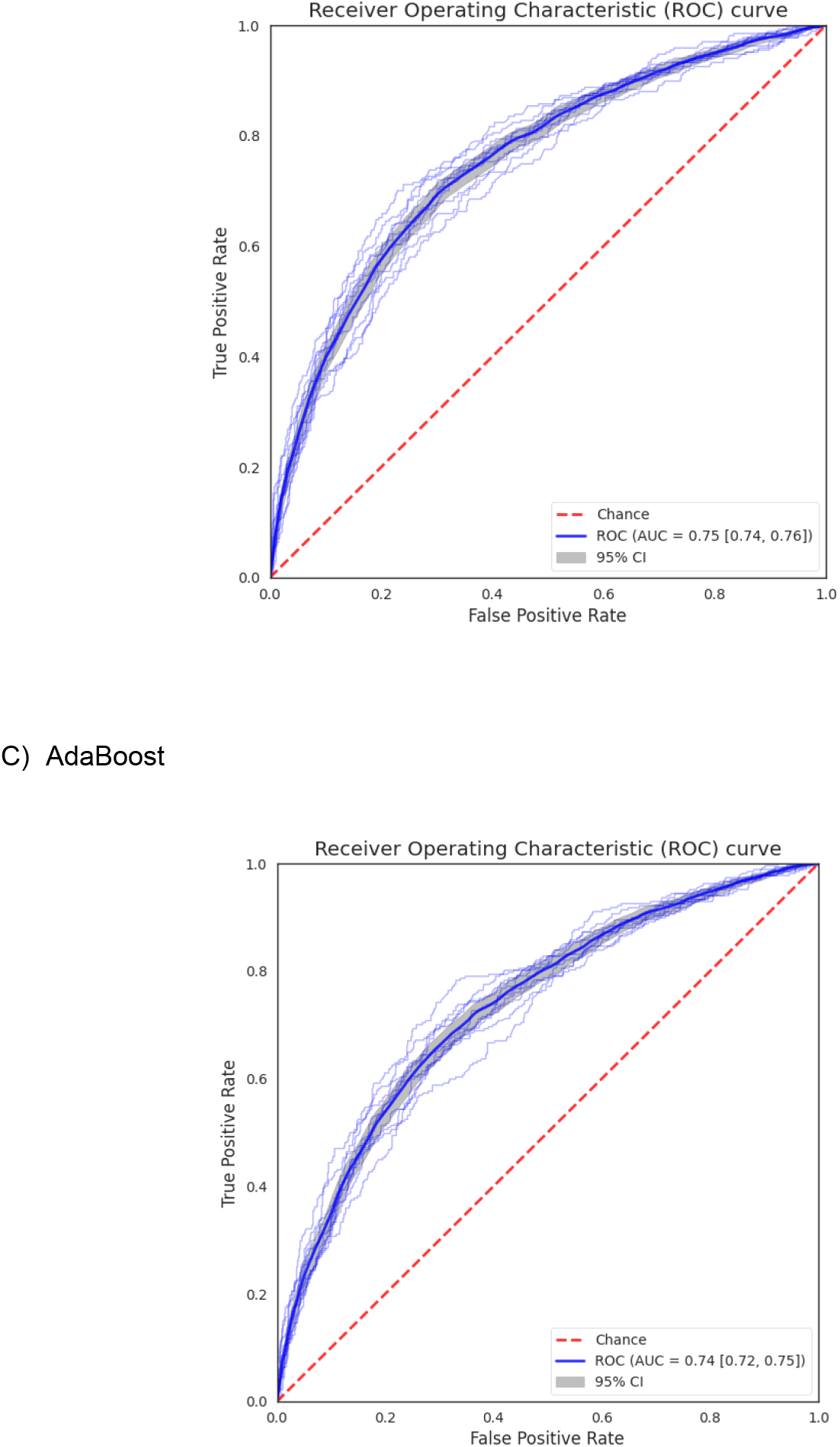
Receiver operating characteristic curve (ROC) of (A) Random Forest, (B) Support Vector Machine and (C) AdaBoost when accounting for the contact with COVID-19 case information. Transparent band shows the 95% Confidence Intervals (CI). The area under the curve (AUC) is provided to summarize the curve.

**Supplementary Figure 4.**
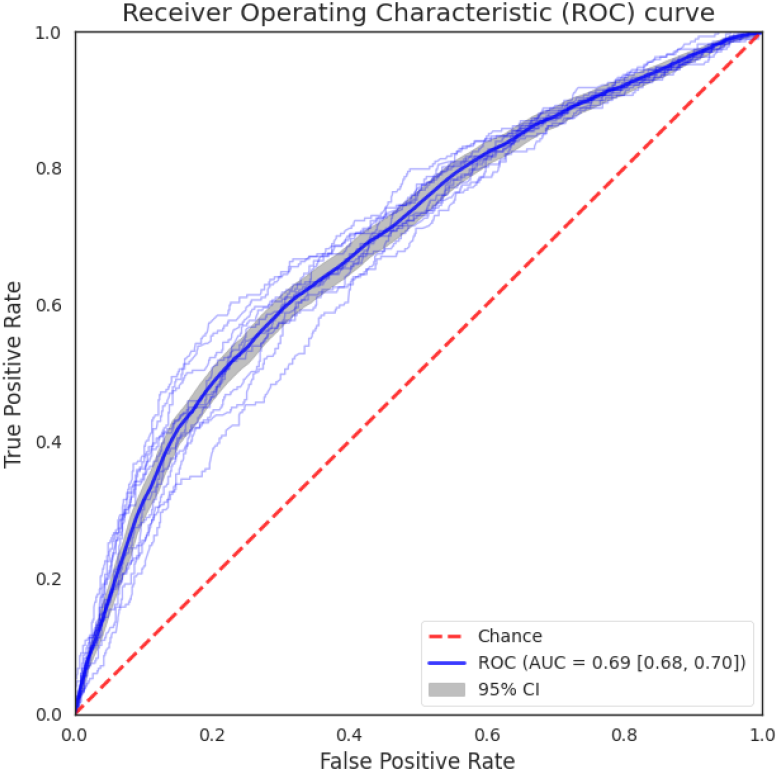
Receiver operating characteristic curve (ROC) of the predictive model when not accounting for the contact with COVID-19 case information. Transparent band shows the 95% Confidence Intervals (CI). The area under the curve (AUC) is provided to summarize the curve.

**Supplementary Figure 5.**
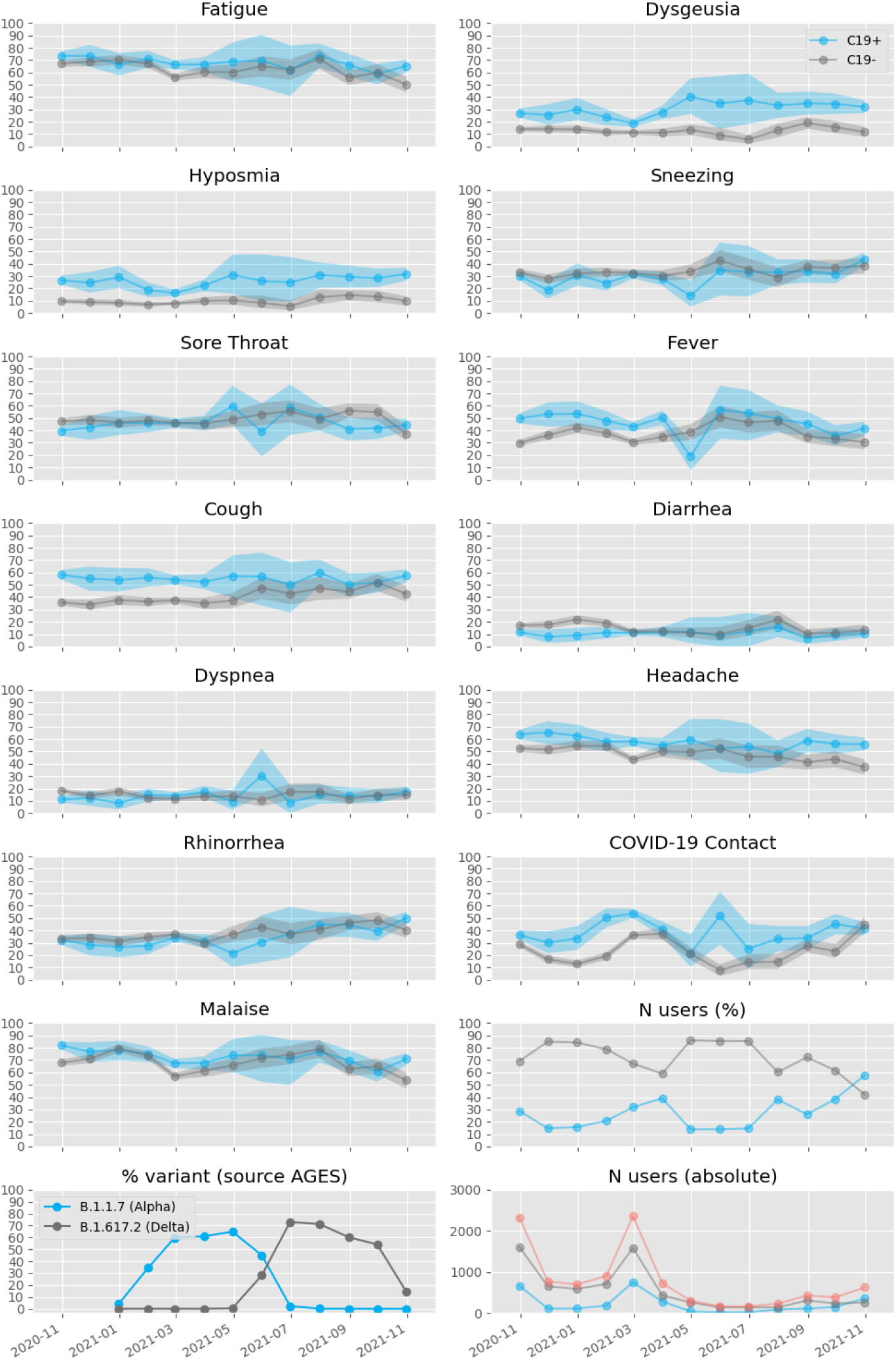
Evolution of the symptom frequencies, the percentage of the Alpha and Delta variants detected in Austria and the number of users.

